# A Novel Arabic-Language Questionnaire to Assess Patient Satisfaction After Refractive Surgery

**DOI:** 10.64898/2026.03.18.26348750

**Authors:** Mortada Abozaid

**Affiliations:** Department of Ophthalmology, Faculty of medicine, Sohag university

**Keywords:** Arabic language, patient satisfaction, questionnaire, refractive surgery

## Abstract

**Purpose:** to develop a questionnaire in Arabic language for assessment of patient satisfaction following refractive surgery.

**Design:** Cross-sectional observational study by survey.

**Patients and methods:** a 25-item questionnaire was self-answered by 327 patients 3 months after having a corneal refractive surgery. The questionnaire was subdivided into 3 subscales: 1-questions to assess vision (15 items), 2-questions to assess eye symptoms (5 items) and 3-questions to assess satisfaction (5 items). Validity and reliability of the questionnaire were assessed using both traditional method by factor analysis and Cronbach’s alpha in addition to Rasch analysis.

**Results:** The preoperative refraction of cases were myopia and myopic astigmatism (297), hyperopia and hyperopic astigmatism (3) and mixed astigmatism (27). Factor loadings ranged from 0.44 to 0.88. Cronbach’s alpha values were 0.894 for the overall scale, 0.858 for the vision subscale, 0.699 for the eye symptoms subscale and 0.720 for the satisfaction subscale. Rasch analysis demonstrated high item reliability (0.94) and person reliability (0.88). Item fit statistics remained mostly within the acceptable range of 0.6 to 1.4.

**Conclusion:** This novel questionnaire is valid and reliable for assessing satisfaction of the patients who had refractive surgery and whose native language is Arabic.

## Introduction

Refr**a**ctive surgery is gaining popularity all over the world due to high success rates, rapid rehabilitation, advances in techniques and platforms and improved safety. At the same time the prevalence of refractive errors is increasing especially myopia which is anticipated to affect about 5 billions (52%) of the world population by 2050. For all these reasons, refractive surgery is becoming one of the most common eye surgeries with estimated 3.6 to 5.8 million precedures per year that is leaded primarly by laser-assisted in situ keratomilleusis (Lasik) and photorefractive keratectomy (PRK) ^(1-3)^.

Unfortunately, the results of refractive surgery are sometimes disappointing to the patients despite good or even excellent objective measures such as postoperative refraction and visual acuity. This makes assessing subjective satisfaction via questionnaires an essential practice that should be done routinely in all patients. This helps to capture complaints that cannot be assessed by ordinary tests which may be too insensitive to them. In addition, questionnaires are considered an essential research tool especially when testing a new maneuver or apparatus or when comparing different techniques in different populations ^(4,5)^.

In Egypt and most countries of the middle east, the native language is Arabic and many patients do not have a second language. At the same time, using a translated version of another language questionnaire would be inappropriate due to environmental and cultural differences. For these reasons, a questionnaire that is primarily designed in Arabic is needed in refractive surgery patients of this region ^(6)^.

The process of developing a new questionnaire is complex and lengthy and involves several steps that should be followed by assessment of its validity and reliability ^(7-10)^. The aim of this study is to develop and validate a new questionnaire in Arabic language that can be used to evaluate satisfaction of patients after refractive surgery.

### Patients and methods

This cross sectional observational study was done using survey via questionnaire followed the tenets of Helsinki declaration and an institutional approval was obtained from the ethics committee of Sohag faculty of medicine. In addition, a written informed consent was obtained from all patients after the nature and aim of the study was explained.

The study included patients 3 months after having an uncomplicated corneal refractive surgery for correction of myopia, hyperopia or mixed astigmatism with age range from 21 to 40 years. The exclusion criteria included patients with previous non refractive eye surgery and those with ocular or systemic diseases or drugs that may affect vision.

A preliminary 50-item questionnaire was designed in Arabic language depending on review of literature, previous validated questionnaires and patients complaints following refractive surgery. Then item reduction to 25 items was done depending on expert focus group discussions.

The 25 items of the questionnaire were divided into three subscales; to assess the vision (15 items), to assess the eye symptoms (5 items) and to assess the satisfaction directly (5 items).

A Lickert scale of 3 choices was used for the responses of each item taking care that the first choice represents the highest satisfaction (given score 2), the second one moderate satisfaction (score 1) and the third one the least satisfaction (score 0) in addition to a fourth choice being (I do not know) in all items.

The overall satisfaction score was calculated as a percentage by summating the total scores of all items answered by a choice other than (I do not know) multiplied by 2 (in the case that all 25 items answered) and multiplied by 2.08, 2.17, 2.27 or 2.38 in cases of 24, 23, 22 or 21 items answered respectively. Patients who answer more than 5 items by (I do not know) are excluded. The overall satisfaction was graded as high (above 80%), moderate (60% to 80%), and low (less than 60%).

The patients included in this study (327 patients and 612 eyes) were divided according to their refractive error into 3 groups; myopia and myopic astigmatism (297 patients), Hyperopia and hyperopic astigmatism (3 patients) and mixed astigmatism (27 patients) and are divided according to the type of refractive surgery into 6 groups; conventional lasik (120 patients), conventional PRK (33), conventional femtolasik (42), wavefront-guided lasik (57), WFG-PRK (63) and WFG femtolasik (12).

The sample size was determined based on established psychometric recommendations for Exploratory Factor Analysis (EFA). A subject-to-item ratio of 10:1 was targeted, which is widely considered the gold standard for providing sufficient statistical power and reliable factor solutions in clinical validation studies. Given the 25 distinct items assessing Visual Function, Ocular Symptoms, and Satisfaction, a minimum sample of 250 participants was sought to ensure that the Principal Component Analysis (PCA) with Varimax rotation would yield stable factor loadings. We collected 327 cases to enhance power of the study.

#### Statistical analysis

The collected data had been coded and verified before computerized data entry. Then data was statistically analyzed using the Statistical Package for the Social Science (SPSS) version 26 and expressed in tables and charts. Construct validity was assessed using factor analysis while Cronbach’s Alpha was used to assess internal consistency or reliability. Rasch analysis ^(11)^ was done using Winsteps (version 16.9, Chicago, IL). The Chi-squared test was used to correlate the satisfaction of patients with their background variables. In all analyses, P < 0.05 indicated statistical significance.

## Results

This study included 327 patients who had refractive surgery for myopia, hyperopia and mixed astigmatism. They responded to the self-administerd questionnaire 3 months after surgery. The mean age of the cases was 26.7±5.6 years with 57 males and 270 females. 285 cases had bilateral surgery and 42 had unilateral surgery (18 right eyes and 24 left eyes).

The group with myopia and myopic astigmatism (297 cases) had a preoperative mean spherical error of -3.50±1.9 D, a mean cylindrical error of -1.39±1.1 and a best-corrected visual acuity of 6/6 in 204 cases (67%) and a postoperative mean spherical error of 0.2±0.6, a mean cylindrical error of -0.5±0.3 and an uncorrected visual acuity of 6/6 in 191 cases (64%). The group with hyperopia and hyperopic astigmatism (3 cases) had a preoperative mean spherical error of +3.25±1.5 D, a mean cylindrical error of -1.64±1.3 and a best-corrected visual acuity of 6/6 in all cases and a postoperative mean spherical error of 0.5±0.8, a mean cylindrical error of -0.30±0.4 and an uncorrected visual acuity of 6/6 in all cases. The group with mixed astigmatism (27 patients) had a preoperative mean spherical error of 1.8±1.4, a mean minus cylindrical error of -3.60±1.8 and a best-corrected visual acuity of 6/6 in 18 cases (66%) and a postoperative mean spherical error of 0.15±0.48, a mean cylindrical error of -0.8±0.89 and an uncorrected visual acuity of 6/6 in 15 cases (55%).

The items which received the majority of answers with (I do not know) are those asking about driving or practicing sports which highlights the importance of the cultural and socioeconomic factors in designing a questionnaire. This can be explained by the fact that most of cases were females who have less access to driving or sport practicing.

### Factor Analysis (Construct Validity)

To validate the three-scale structure, Exploratory Factor Analysis (EFA) was performed using the Principal Component Analysis (PCA) extraction method.

Suitability: Data suitability was confirmed via the Kaiser-Meyer-Olkin (KMO) measure of sampling adequacy and Bartlett’s test of sphericity to ensure the correlation matrix was not an identity matrix.

Rotation: A Varimax rotation with Kaiser Normalization was applied to maximize the variance of the squared loadings, ensuring each question loaded clearly onto one of the three intended factors.

Retention Criteria: Factors were retained based on Eigenvalues > 1.0 and inspection of the Scree Plot. Items with factor loadings < 0.4 were considered for revision and the current items demonstrate strong alignment with their respective categories as shown in Table 1.

**Table 1:** Factor loadings of the 25 questions:

| Item | Factor 1: Visual<br>Function | Factor 2: Ocular<br>Symptoms | Factor 3: Satisfaction |
| --- | --- | --- | --- |
| Q1: Day Vision | 0.53 | 0.12 | 0.09 |
| Q2: Night Vision | 0.65 | 0.15 | 0.11 |
| Q3: Reading/Subtitles | 0.53 | 0.08 | 0.21 |
| Q4: Smart Phones | 0.65 | 0.22 | 0.14 |
| Q5: Playing sports | 0.68 | 0.11 | 0.18 |
| Q6: Near vision | 0.68 | 0.09 | 0.22 |
| Q7: Reading/Studying | 0.69 | 0.19 | 0.18 |
| Q 8: Vision immediately after awakening | 0.58 | 0.24 | 0.12 |
| Q9: Night driving | 0.61 | 0.14 | 0.10 |
| Q10: Halos around lights | 0.55 | 0.31 | 0.08 |
| Q11: Fluctuation of vision | 0.52 | 0.28 | 0.14 |
| Q12: Difference in the 2 eyes vision | 0.48 | 0.19 | 0.25 |
| Q13: Double vision | 0.44 | 0.21 | 0.19 |
| Q14: Color vision | 0.61 | 0.06 | 0.28 |
| Q15: Judging Distance | 0.64 | 0.05 | 0.25 |
| Q16: Burning Sensation | 0.14 | 0.88 | 0.06 |
| Q17: Foreign Body Sensation | 0.09 | 0.85 | 0.12 |
| Q18: Light Sensitivity | 0.21 | 0.79 | 0.10 |
| Q19: Watering of Eye | 0.18 | 0.74 | 0.08 |
| Q20: Headache | 0.05 | 0.68 | 0.15 |
| Q21: Worry/Deterioration | 0.12 | 0.11 | 0.89 |
| Q22: Need for Glasses | 0.22 | 0.09 | 0.84 |
| Q23:Self-sufficiency after surgery | 0.35 | 0.18 | 0.62 |
| Q24: Satisfaction | 0.31 | 0.14 | 0.81 |
| Q25: Regret | 0.19 | 0.07 | 0.78 |

Factor analysis performed on all items showed that the 3 subscales were consistant

#### Reliability Analysis

Internal consistency for the overall scale and each of the three subscales was calculated using Cronbach’s Alpha (alpha) by using the scale of items in SPSS program as shown in table 2 and 3.

**Table 2:**
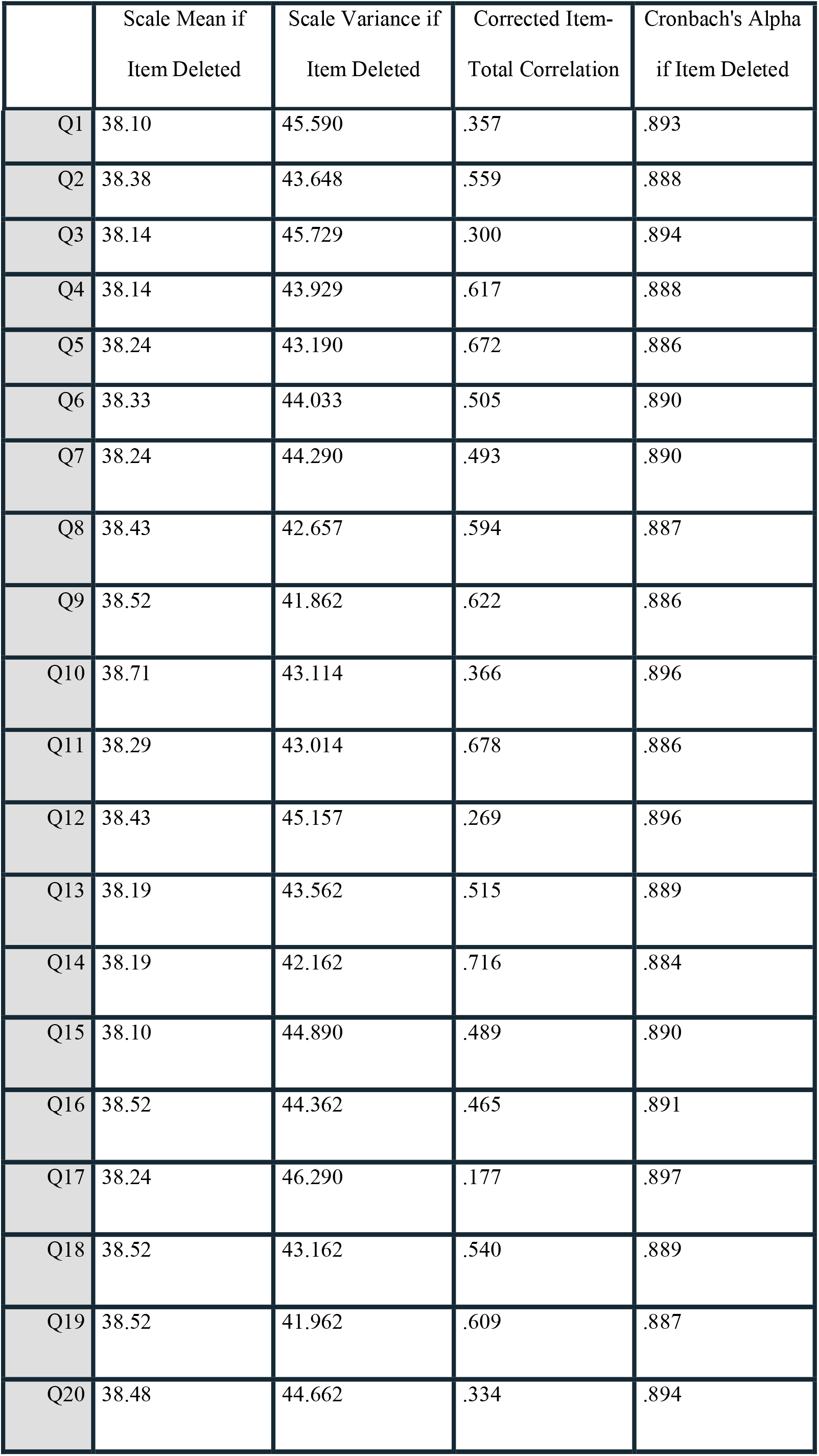
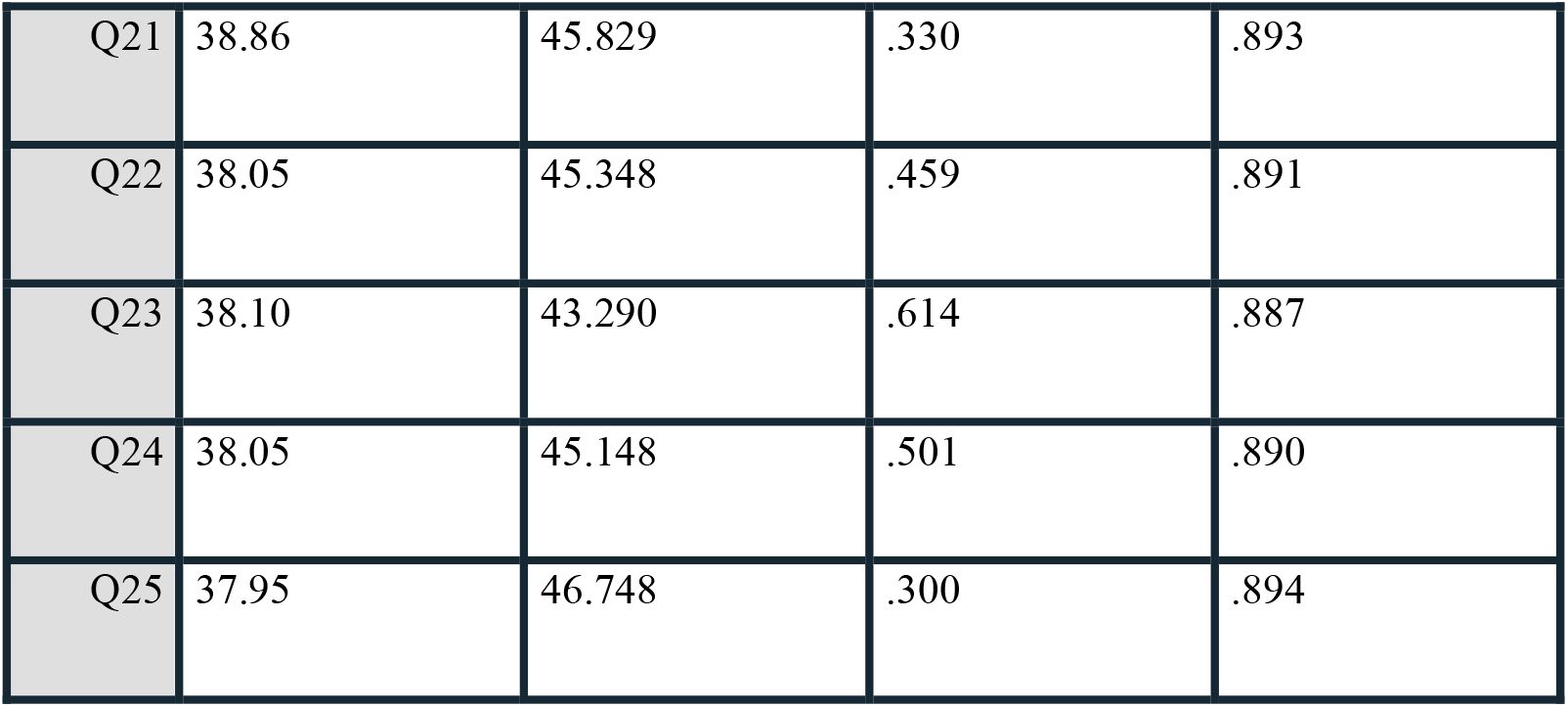
Item-Total Statistics.

**Table 3.**
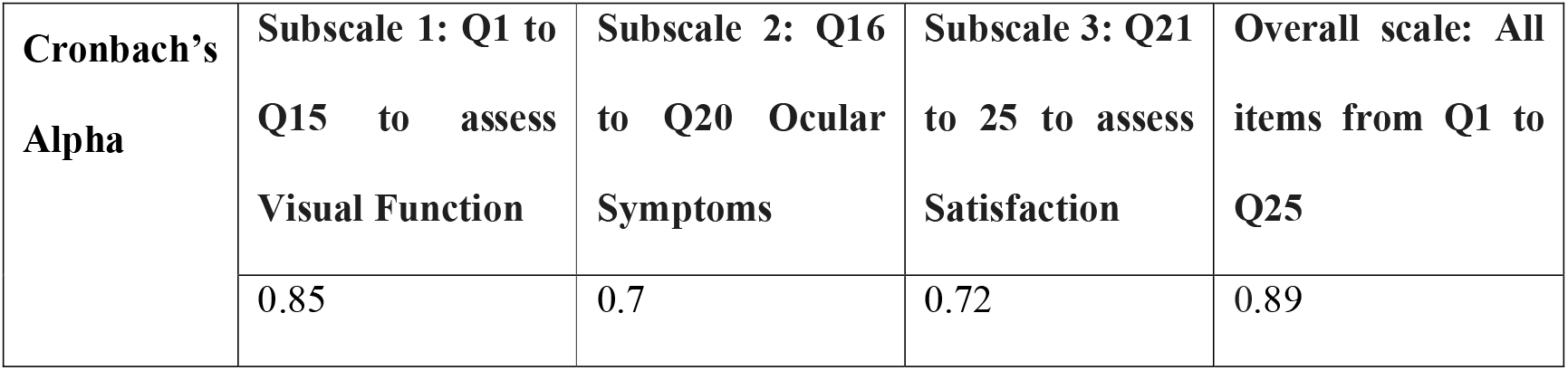
Cronbach’s Alpha values for the overall scale and the 3 subscales.

**Table 4.** Rasch item measures.

| **Item ID** | **Question Description** | **Difficulty Measure (Logit)** | **Infit MNSQ** | **Outfit MNSQ** | **Position on Map** |
| --- | --- | --- | --- | --- | --- |
| **Q9** | Night Driving | +1.85 | 1.12 | 1.25 | **Hardest** |
| **Q10** | Halos around lights | +1.42 | 1.38 | 1.45 | High Difficulty |
| **Q2** | Night Vision | +1.10 | 1.05 | 1.08 | High Difficulty |
| **Q11** | Fluctuation of vision | +0.88 | 1.21 | 1.19 | Above Average |
| **Q13** | Double vision | +0.75 | 0.95 | 0.92 | Above Average |
| **Q12** | Difference in the 2 eyes | +0.62 | 1.10 | 1.15 | Above Average |
| **Q20** | Headache | +0.48 | 1.02 | 1.01 | Moderate |
| **Q18** | Light Sensitivity | +0.35 | 1.08 | 1.12 | Moderate |
| **Q16** | Burning Sensation | +0.22 | 0.98 | 0.95 | Moderate |
| **Q17** | Foreign Body Sensation | +0.15 | 1.04 | 1.03 | Moderate |
| **Q19** | Watering of Eye | +0.08 | 0.96 | 0.94 | **Mean Difficulty** |
| **Q8** | Vision after awakening | -0.12 | 1.15 | 1.10 | Moderate |
| **Q15** | Judging Distance | -0.25 | 0.90 | 0.88 | Moderate |
| **Q3** | Reading/Subtitles | -0.38 | 0.88 | 0.82 | Easy |
| **Q7** | Reading/Studying | -0.45 | 0.85 | 0.80 | Easy |
| **Q25** | Regret (re-operation) | -0.58 | 0.92 | 0.85 | Easy |
| **Q6** | Near vision | -0.65 | 0.94 | 0.89 | Easy |
| **Q4** | Smart Phones | -0.72 | 0.82 | 0.78 | Easy |
| **Q5** | Playing sports | -0.85 | 0.88 | 0.84 | Easy |
| **Q21** | Worry/Deterioration | -0.92 | 0.95 | 0.90 | Easy |
| **Q14** | Color vision | -1.05 | 0.80 | 0.75 | Very Easy |
| **Q24** | Satisfaction | -1.15 | 0.92 | 0.88 | Very Easy |
| **Q23** | Self-sufficiency | -1.45 | 0.85 | 0.80 | Very Easy |
| **Q22** | Need for Glasses | -1.75 | 0.78 | 0.70 | Very Easy |
| **Q1** | Day Vision | -2.10 | 0.85 | 0.72 | **Easiest** |

**Table 5:**
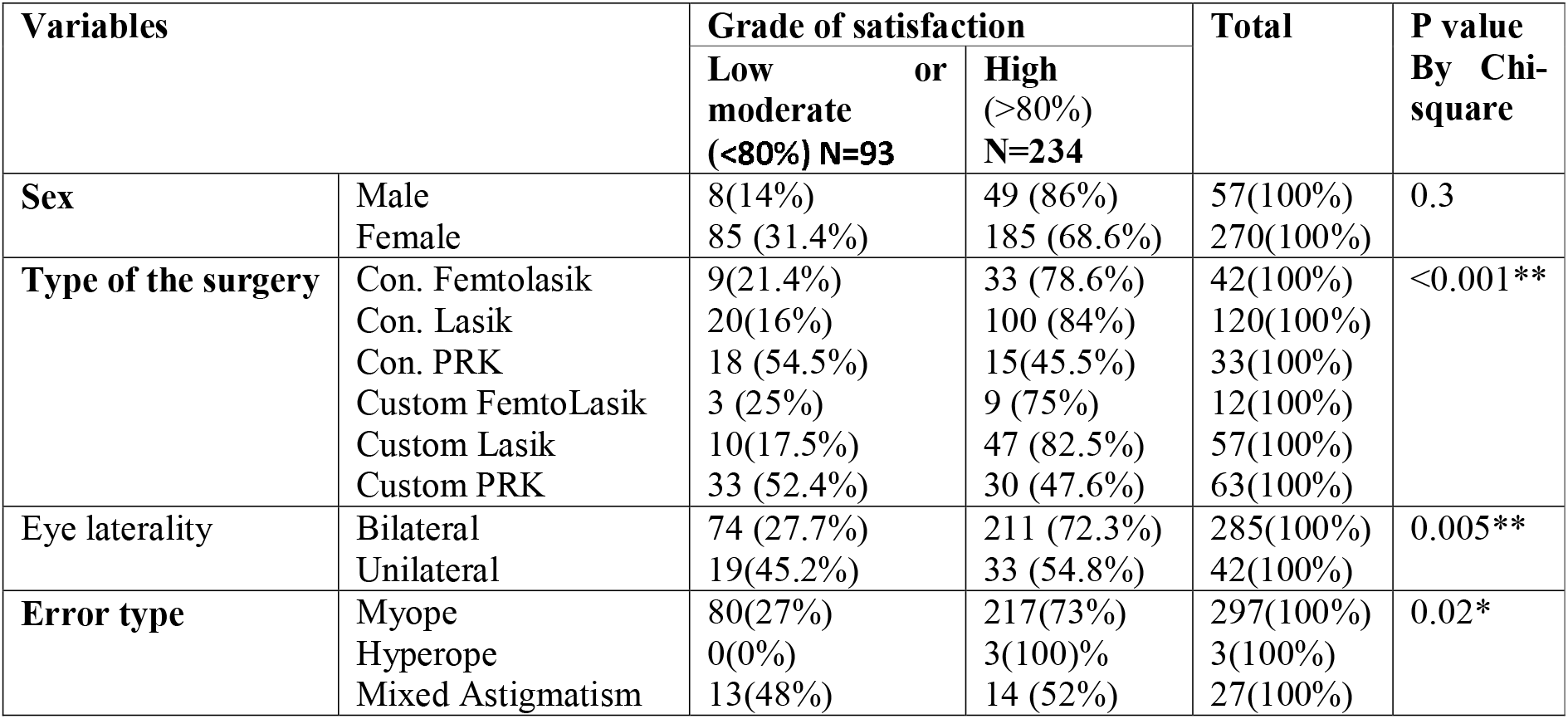
Relation between the degree of satisfaction and sex, type and laterality of the surgery and type of the error of refraction .N=327.

Rasch analysis was conducted to evaluate the psychometric properties of the 25-item instrument using a sample of 327 respondents. The results demonstrated high item reliability (0.94) and person reliability (0.88). Item fit statistics remained mostly within the acceptable range of 0.6 to 1.4, with the exception of Q10 (Halos), which exhibited slight underfit (Outfit MNSQ = 1.45). The Person-Item map revealed that while the items effectively cover a range of visual functions, the sample mean ability was slightly higher than the item mean difficulty, suggesting a potential ceiling effect for patients with high visual recovery. The category thresholds were ordered correctly, supporting the use of the three-point rating scale.

### Factors affecting satisfaction

Considering 80% as the boundery between high and low satisfaction, it is noticed that bilateral surgery, preoperative myopic error and Lasik surgery were associated wither higher satisfaction. On the contrary unilateral surgery, preoperative mixed astigmatism and PRK are associated with lower satisfaction. In addition, sex and customized surgery did not show differences in the level of satisfaction. It is worth mentioning that the lower satisfaction with PRK than Lasik could be due to the short interval between surgery and answering the questionnaire (3 months) when PRK may still in need for longer period to reach complete healing

## Discussion

Questionnaires are increasingly used to assess the quality of vision following different ocular surgeries especially refractive ones. The major advantage of such questionnaires is its ability to pick up the complaints that cannot be assessed objectively and which can markedly affect the patient’s attitude of the surgery even in the presence of good visual acuity and postoperative refraction.

A good questionnaire should be self-administered by the patient, formulated in his native language and considers his culture. In this study a novel questionnaire designed in Arabic language was validated in assessing patient’s satisfaction following corneal refractive surgery.

Brett L Halliday ^(12)^ assessed satisfaction of 108 patients one year after having PRK for myopia. He used a simple 4-item questionnaire with questions about satisfaction with the procedure, glare, distortion and the need for refractive correction. Twenty five patients (24%) made negative comments

Vitale and colleagues ^(13)^ developed and validated the refractive status and vision profile (RSVP) questionnaire to measure vision-related quality of life in patients with refractive errors or who had a refractive surgery. The questionnaire included 42 items and was divided into 8 subscales. Chronbach’s alpha was 0.92 for the overall scale and ranged from 0.70 to 0.90 for the subscales.

Pesudovs et al ^(14)^ developed and validated the quality of life impact of refractive correction (QIRC) using both traditional methods and Rasch analysis to asses quality of life of patients with refractive correction including spectacles, contact lenses and refractive surgery. A 90-item pilot questionnaire was reduced to 20 items which showed good validity and reliability with a factor loading range of 0.40 to 0.76 and a Cronbach’s alpha of o.78 and a Rasch analysis showing good precision, reliability and internal consistency.

Tran and Manche ^(15)^ tried to detect the factors that affect patient satisfaction following refractive surgery using selected items from the National Eye Institute Visual Function Questionnaire-25 and the Press Ganey survey. They recruited 53 patients over 3 years with 89% having Lasik and 11% having PRK and used student’s t test to measure Correlation between patient clinical and sociodemographic variables and questionnaire items. They found that high satisfaction scores were independent of patient-specific characteristics.

This novel Arabic language questionnaire is concise (25 items only) and is designed specifically to measure satisfaction after refractive surgery (Lasik, femtolasik and PRK). Validity and reliability were confirmed using both traditional methods and Rasch analysis. The study showed that high satisfaction was noted in patients with bilateral surgery, preoperative myopic error or lasik surgery.

The major advantage of this study is the development of a questionnaire that is originally prepared in Arabic language with an English version that can be used in other parts of the world. In addition, the method of calculating the scores of satisfaction is simple and easy to explain to patients. While the main limitations are including only corneal refractive surgery while other surgeries such as phakic intraocular lens and clear lens extraction are not tested. In addition, the patients included have mainly myopia and myopic astigmatism with little cases having mixed astigmatism or hyperopia.

In the future, it is planned to transform the questionnaire into a mobile application that can be easily downloaded by patients and doctors, thus spreading the practice of satisfaction assessment to increase understanding of side effects of refractive surgery and finally improving its outcome. In addition, scoring of the questionnaire can be altered so as to make use of Rasch analysis in varying the weight of each item according to its impact on vision and response of the patients.

## Data Availability

All data produced in the present study are available upon reasonable request to the authors

