## Appendix 2 for "A Novel Arabic-Language Questionnaire to Assess Patient Satisfaction After Refractive Surgery"

**A Questionnaire to Asses Satisfaction After Refractive Surgey**

Name:

Sex: 1-male	2-female

Date of birth:

Date of surgery:

Type of surgery: 1-Lasik: 1- conventional 2- Customized

2- Femtolasik: 1- conventional 2- Customized

3- PRK: 1- conventional 2- Customized

Operated eye: 1- right 2- left 3- both

A) **Questions to assess vision:**
1- Day vision: 1- Excellent 2- Good 3- Bad 4- I don’t know
2- Night vision: 1- Excellent 2- Good 3- Bad 4- I don’t know
3- Reading news ticker and movies subtitles: 1- Excellent 2- Good 3- Bad 4- I don’t know
4- Using smart phones: 1- Excellent 2- Good 3- Bad 4- I don’t know
5- Playing sports: 1- Excellent 2- Good 3- Bad 4- I don’t know
6- Near vision such as threading a needle or seeing alarm clock: 1- Excellent 2- Good 3- Bad 4- I don’t know
7- Reading and studying: 1- Excellent 2- Good 3- Bad 4- I don’t know
8- Vision immediately after awakening: 1- Excellent 2- Good 3- Bad 4- I don’t know
9- Night driving: 1- Excellent 2- Good 3- Bad 4- I don’t know
10- Halos around lights: 1- Non 2-mild 3- severe 4- I don’t know
11- Fluctuation of vision: 1- Non 2-mild 3- severe 4- I don’t know
12- Difference in vision between the 2 eyes: 1- Non 2-mild 3- severe 4- I don’t know
13- Double vision: 1- Non 2-mild 3- severe 4- I don’t know
14- Color vision: 1- Excellent 2- Good 3- Bad 4- I don’t know
15- Judging distance while going upstairs or downstairs: 1- Excellent 2- Good 3- Bad 4- I don’t know
B) **Questions to assess eye symptoms:**
16- Burning sensation in the eye: 1- Non 2-mild 3- severe 4- I don’t know
17- Foreign body sensation in the eye: 1- Non 2-mild 3- severe 4- I don’t know
18- Light sensitivity: 1- Non 2-mild 3- severe 4- I don’t know
19- Watering of the eye: 1- Non 2-mild 3- severe 4- I don’t know
20- Headache: 1- Non 2-mild 3- severe 4- I don’t know
C) **Questions to assess satisfaction after surgery:**
21- Worry about deterioration of vision: 1- Non 2-mild 3- severe 4- I don’t know
22- Need for glasses after surgery: 1- Non 2-sometimes 3- always 4- I don’t know
23- Self sufficiency after surgery: 1- Increased 2- Same 3- Less 4- I don’t know
24- Satisfaction about surgery: 1- Excellent 2- Good 3- Bad 4- I don’t know
25- Do you regret to do the surgery: 1- Never 2-Sometimes 3- Always 4- I don’t know
