## Appendix 1 for "A Novel Arabic-Language Questionnaire to Assess Patient Satisfaction After Refractive Surgery"

**استبيان لقياس مستوي الرضا بعد عملية تصحيح الابصار**

**الاسم:**

**النوع**: 1- ذكر	2-انثي

**تاريخ الميلاد:**

**تاريخ اجراء العملية:**

**نوع العملية:**

1**- ليزك**: 1-عادى 2- تفصيل

**2- فمتوليزك**: 1-عادى 2- تفصيل

3**- ليزر سطحى (بي ار كيه):** 1-عادى 2- تفصيل

**العين:** 1-اليمني	2-اليسرى	3-العينين

1. **أسئلة لقياس مستوى النظر:**
1- **الرؤية بالنهار:** 1-ممتازة 2- جيدة 3- سيئة 4- لا أعرف
2- **الرؤية بالليل:** 1-ممتازة 2- جيدة 3- سيئة 4- لا أعرف
3- **قراءة شريط الاخبارأو ترجمة الأفلام الأجنبية:** 1-ممتازة 2- جيدة 3- سيئة 4- لا أعرف
4- **استعمال الهاتف المحمول:** 1-ممتاز 2- جيد 3- سيئ 4- لا أعرف
5- **ممارسة الرياضة:** 1-ممتازة 2- جيدة 3- سيئة 4- لا أعرف
6- **رؤية الاشياء القريبة مثل لضم الابرة أو المنبه:** 1-ممتازة 2- جيدة 3- سيئة 4- لا أعرف
7- **المذاكرة والقراءة:** 1-ممتازة 2- جيدة 3- سيئة 4- لا أعرف
8- **الرؤية بعد الاستيقاظ مباشرة:** 1-ممتازة 2- جيدة 3- سيئة 4- لا أعرف
9- **القيادة الليلية:** 1-ممتازة 2- جيدة 3- سيئة 4- لا أعرف
10- **هالات مضيئة:** 1-لا يوجد 2- هالات قليلة 3- هالات عديدة 4- لا أعرف
11- **تذبذب في الرؤية اثناء اليوم:** 1-لا يوجد 2- تذبذب بسيط 3- تذبذب شديد 4- لا أعرف
12- **اختلاف فى النظر بين العينين:** 1-لا يوجد 2- اختلاف بسيط 3- أختلاف شديد 4- لا أعرف
13- **ازدواج في الرؤية:** 1-لا يوجد 2- ازدواج بسيط 3- ازدواج شديد 4- لا أعرف
14- **تمييزالألوان:** 1-ممتاز 2- جيد 3- سيئ 4- لا أعرف
15- **تحديد درجات السلم اثناء الطلوع والنزول:** 1-ممتاز 2- جيد 3- سيئ 4- لا أعرف
17. **أسئلة لقياس وجود أعراض مرضية في العين:**
16- **الشعور بحرقان في العين:** 1-لا يوجد 2- الم بسيط 3- ألم شديد 4- لا أعرف
17- **الشعور بوجود جسم غريب او رمل داخل العين**: 1-لا يوجد 2- شعور بسيط 3- شعور شديد 4- لا أعرف
18- **وجود حساسية للضوء:** 1-لا يوجد 2- حساسية بسيطة 3- حساسية شديدة 4- لا أعرف
19- **تدميع العين:** 1-لا يوجد 2- تدميع بسيط 3- تدميع شديد 4- لا أعرف
20- **صداع**: -لا يوجد 2- صداع بسيط 3- صداع شديد 4- لا أعرف

**ج) أسئلة لقياس مستوى الرضا عن العملية:**

21- **القلق من ضعف النظر مرة اخرى:** 1-لا يوجد 2- قلق بسيط 3- قلق شديد 4- لا أعرف
22- **الحاجة الى ارتداء نظارة بعد العملية:** 1- لا يوجد 2- أحيانا 3- حاجة دائمة 4- لا أعرف
23- **درجة الثقة بالنفس بعد العملية:** 1- كبيرة 2- متوسطة 3- منخفضة 4- لا أعرف
24- **درجة الرضا عن النظر بعد العملية:** 1- ممتازة 2- عادية 3- غير راضى 4- لا أعرف
25- **هل انت نادم علي اجراء العملية**: 1- اطلاقا 2- احيانا 3- دائما 4- لا أعرف
